# Lung ultrasound to assess pulmonary congestion in patients with acute exacerbation of COPD - a feasibility study

**DOI:** 10.1101/2022.07.28.22277514

**Authors:** Øyvind Johannessen, Fride Uthaug Reite, Rahul Bhatnagar, Tarjei Øvrebotten, Gunnar Einvik, Peder L. Myhre

## Abstract

**Background:** Chronic heart failure (HF) coexist with chronic obstructive pulmonary disease (COPD) in approximately 25% of patients and is associated with worse outcomes. Lung ultrasound (LUS) is a validated technique to diagnose pulmonary congestion by detecting vertical lung artifacts, B-lines. Pulmonary inflammation is also associated with B-lines, but little is known about LUS in patients with acute exacerbation of COPD (AECOPD).

**Aims:** To assess the feasibility of LUS to detect concurrent acute HF in AECOPD and examine the associations between B-lines, clinical parameters during hospitalization and re-hospitalizations and mortality.

**Methods & results:** In a prospective cohort study 123 patients with AECOPD (age 75±9 years, 57 [46%] men) underwent 8-zone bedside LUS within 24h after admission. A positive LUS was defined by ≥3 B-lines in ≥2 zones bilaterally. A cardiologist committee blinded for LUS adjudicated whether concurrent HF was present (n=48, 39%). The median number of B-lines was 8 (IQR 5-13) and 16 (13%) patients had positive LUS. Positive LUS was associated with infiltrates on chest X-ray. The prevalence of positive LUS was similar with and without concurrent HF 8 (17%) vs 8 (11%), p=0.34, while the number of B-lines was higher in concurrent HF: median 10 (IQR 6–16) vs 7 (IQR 5-12) (p=0.03). The sensitivity and specificity for positive LUS to detect concurrent HF was 16.7% and 89.3%, respectively. Positive LUS was not associated with re-hospitalization and mortality: Adjusted HR 0.93 (0.49-1.75), p=0.81.

**Conclusions:** LUS did not detect concurrent HF or predict risk in patients hospitalized with AECOPD.

## Introduction

Chronic obstructive pulmonary disease (COPD) and chronic heart failure (CHF) are increasing global epidemics, estimated to affect approximately 175 and 26 million people worldwide, respectively.^1,2^ These diseases share risk factors and symptoms and both propose major public health challenges due to substantial morbidity and mortality.^3^ It is estimated that approximately 25% of COPD patients have coexisting CHF, and the combination is associated with worse outcomes.^4^ While current treatment options for improving outcome in COPD are limited, potent life-saving medications for CHF exists. Detecting and treating CHF in COPD is therefore important.

Lung ultrasound (LUS) is a quick, easy and validated technique used to diagnose and assess pulmonary congestion in patients with suspected acute HF in the emergency department.^5,6^ Current guidelines recommend its use in the initial diagnostic work-up of acute HF.^2^ LUS relies on detection of pleural effusions and vertical artifacts, B-lines. B-lines demonstrates high sensitivity and specificity for extravascular lung water, a major property of congestive HF.^7^ B-lines on LUS may also indicate pathophysiological processes associated with increased lung density, i.e. lung parenchyma inflammation. Increased lung density is not one of the hallmarks of COPD, thus, LUS has the potential to discriminate HF from non-parenchymal lung disease like COPD.^5,6,8^ However, the current empirical evidences for LUS are derived from studies of patients with suspected acute HF or undifferentiated patients with acute dyspnea and the ability of LUS to discriminate concurrent acute HF in patients with AECOPD is unclear.^9-11^

In this study bedside LUS was performed in patients with AECOPD to 1) assess the feasibility of LUS to detect concurrent acute HF in AECOPD and 2) examine the associations between B-lines, clinical parameters during hospitalization and re-hospitalizations and mortality.

## Methods

### Patient population

We conducted a prospective cohort study at Akershus University Hospital between May and June 2017, and between February 2020 and September 2021. Patients ≥ 18 years, admitted with a tentative diagnosis of AECOPD, were assessed for eligibility at the morning rounds at days were the trained LUS-technicians (ØJ, FU) were present. We excluded patients with pneumothorax, thorax deformities, recent thoracic surgery, comorbid asthma, pulmonary fibrosis or pleural disease, and COVID-19, as these conditions may interfere with the interpretation of B-lines.^8^ Other exclusion criteria were mental disorientation, organic delirium, dementia, or other factors that hinder obtaining informed consent. Baseline clinical characteristics, prior co-morbidities, regular medications, and laboratory data were extracted from the hospital electronic health records.

The study was conducted in accordance with the Declaration of Helsinki and was approved by the regional medical ethics committee (REK approval no. 2017/663) and by the local Data protection officer (project approval 17/135). In cases where LUS uncovered pathological conditions not acknowledged by the attending physician the attending physician was notified and prompted to consider relevant investigations.

### Lung ultrasound protocol and B-line analysis

The examinations were performed daily, except on weekends, for three consecutive days. The examination protocol was according to international guidelines for LUS to assess pulmonary congestion.^5,8,12^ All LUS examinations were done with two types of low-cost handheld ultrasound devices. We examined the first group (n=60) with a dual-probe V-scan Extend™ (General Electric, Vingmed, Horten, Norway). For this first patient group, a linear array transducer was used with bandwidth 3.3–8.0 MHz. The scanning depth was set to 5 cm, and clips of 3 seconds were recorded in each lung zone. The second group (n=65) were examined with a Phillips Lumify™, (Phillips) using an S4-1, broadband sector array transducer (bandwidth 1-4 MHz) using the cardiac setting. We recorded 6-second cine-loops in 8 thoracic zones and transferred the cine loops to a computer for offline storage and analysis. The images/cine-loops were de-identified and read by a single examinator (ØJ). Temporal and clinical blinding was assured before image analysis.

LUS B-lines were defined as discrete hyperechoic vertical artifacts arising from the pleural line and stretching from the pleural line to the edge of the screen. To quantify B-lines, we counted and summed the highest number of B-lines from a freeze-frame after reviewing the entire clip from each of the 8 lung zones. This approach was adopted from prior LUS-studies and provided the basis for the primary LUS analysis.^13-15^ Patients with ≥3 missing lung zones on either side were excluded from further analysis (n=1). Missing data from lung zones were imputed if there were no more than two missing zones per examination. The mean number of B-lines from adjacent valid zones were imputed to the missing lung zones to obtain a full dataset with 123 patients (**Supplemental Material)**. The patients reported perceived strenuousness on a 5-point Likert scale. Two investigators (OJ and FU) performed all examinations and measurements, and inter-rater agreement were assessed in 10 consecutive patients. Temporal blinding was assured during measurements.

In accordance with multiple previous studies the presence of three or more B-lines in two or more thoracic zones bilaterally was used to define a ‘positive LUS’, i.e. alveolar interstitial syndrome (AIS) suggestive of pulmonary congestion.^8^

### Clinical outcomes

A clinical endpoint committee (CEC) composed of three cardiologists, (RB, TØ and PLM), blinded to all LUS findings, independently reviewed each case to adjudicate 1) HF presence and a concurrent cause for exacerbation, 2) HF presence, but not a concurrent cause of exacerbation and 3) HF not present. The CEC had full access to the patient records, including charts, laboratory parameters, imaging (chest x-rays and computed tomography), electrocardiograms and echocardiograms, and adjudicated according to ESC HF guidelines.^16^

A composite outcome of rehospitalization for AECOPD and all-cause mortality within 12 months after enrollment was used as the primary clinical outcome. Outcome data were obtained through review of the patients electronic health records and a hospitalization for AECOPD was defined as a final diagnosis of AECOPD and objective findings, typical history and treatment for COPD.

### Other clinical data

All patients underwent routine clinical diagnostic work-up for acute dyspnea on hospital admission in the emergency department (ED) before admission to the pulmonary deprtment. The work-up included clinical examination including height and weigth, standard laboratory values (creatinin, C-reactive protein [CRP]), blood gas analysis and chest X-ray. In addition, N-terminal pro-B-type natriuretic peptide (NT-proBNP) concentration and high-sensitivity cardiac Troponin T (cTnT) (both Roche Diagnostics, Basel Switzerland) were sampled at the clinician’s discretion. The last known spirometry result, including forced expiratory volume during 1^st^ second (FEV_1_) was obtained from the medical records. Presence of pulmonary congestion or an infiltrate as described by the radiologist in clinical routine was registered. Point of care LUS was not implemented as standard work-up of acute dyspena in the ED.

### Statistical analysis

Continuous data are expressed as means ± standard deviation (SD), or median with interquartile range (IQR). Categorical data are expressed as counts and percentages. Between-group comparisons of baseline characteristics were analyzed with two sample t-test, Wilcoxon rank sum test or Chi-square, as appropriate. Skewed variables (NT-proBNP, CRP, and cTnT) were log-transformed prior to regression analysis.

The diagnostic properties of a positive LUS to correctly classify concurrent HF in AECOPD was assessed using receiver operator characteristics (ROC) analysis. We also calculated the sensitivity and specificity. We performed *a priori* sensitivity analysis, excluding patients with HF present but not as a concurrent cause of the exacerbation. The number of B-lines as continuous variable across 8 zones was also used for assessing the clinical endpoints of concurrent HF. And we also explored a less strict threshold for a positive LUS (≥3 B-lines in one zone bilaterally).

We used univariable logistic regression to explore the associations between positive LUS examination and the following clinical variables: Age, sex, body mass index (BMI), smoking, log transformed NT-proBNP, log transformed CRP, log transformed creatinine, partial pressure of oxygen, diuretic treatment before LUS examination, congestion on CXR, and infiltrate on CXR.

B-lines are discrete count variables and we used unadjusted and adjusted negative binomial regression models (NBR) to explore associations between the total sum of B-lines and variables known to be associated with B-lines: Age, BMI, diuretics before LUS, congestion on CXR and infiltrate on CXR.^17-21^ Results from the NBR models are reported as ratios with 95% confidence intervals (CI) and represent % increase or decrease in B-lines. Dynamic change in B-lines between consecutive LUS were assessed with the Wilcoxon signed rank test.

Kaplan-Meier plots for the cumulative rate of AECOPD rehospitalizations or all-cause death for the positive and negative LUS examination were developed. We used Cox proportional hazard models to analyze the association between positive LUS and time to AECOPD rehospitalization or all-cause death, adjusting for clinical risk factors known to be associated with adverse outcomes in patients hospitalized for AECOPD: age, sex, and FEV_1_.

Statistical software Stata SE version 17.0 (Statacorp. College station, Texas, USA 2015) was used for all data calculations. A double-sided p-value of <0.05 was considered significant.

## Results

### Study population and patient characteristics

In total, 123 patients (mean age 75 ± 9 years, 57 (46%) men) with AECOPD underwent LUS (n=60 in the first and n=63 in the second inclusion period) (**Figure 1**). Valid data was obtained from all zones in 116 (94%) and we imputed missing zones in 7 (6%) patients. The median B-line number in LUS was 8 (IQR 5-13, range 0 to 33) at baseline. At successive scans the median number of B-lines was 8 (IQR 5-12, range 0 to 21) at day 2 (n=38), with no significant change from baseline (p=0.07). At day 3 (n=19) the median number of B-lines was 7 (IQR 1-11, range 0 to 23).

**Figure 1.**
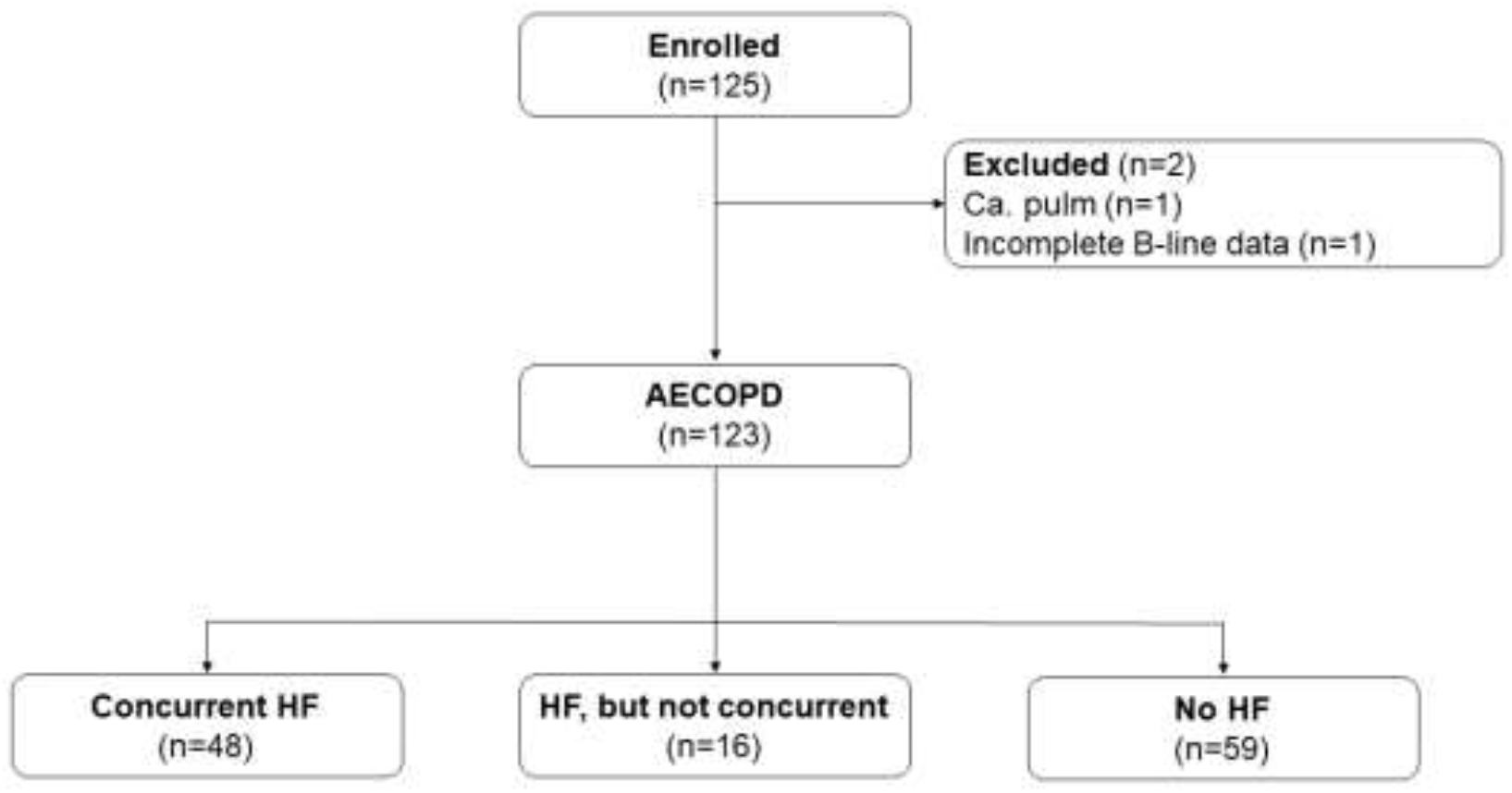
Flow chart AECOPD=acute exacerbation of chronic obstructive pulmonary disease, HF=heart failure

The majority (75%) of the patients had moderate or severe COPD (**Table 1**) and all patients received standard AECOPD treatment with bronchodilators, systemic corticosteroids, and antibiotics at the clinicians’ discretion. The median length of hospital stay was 5 (IQR 3-5) days. Between admission and LUS examination, 17 (14%) patients initiated diuretic treatment.

**Table 1.** Baseline characteristics of the total population, and by concurrent HF. Mean (standard deviation), n (%) or median (25-75^th^ percentile).

|  | Total | No concurrent HF | HF concurrent | p-value |
| --- | --- | --- | --- | --- |
|  | *N=123* | *N=75* | *N=48* |  |
| Age (years) | 75 ± 9 | 72 ± 9 | 78 ± 8 | <0.001 |
| Male | 57 (46%) | 31 (41%) | 26 (54%) | 0.16 |
| Smoking | 52 (42%) | 34 (45%) | 18 (38%) | 0.39 |
| Body mass index (kg/cm2) | 23.8 ± 6.4 | 23.1 ± 5.1 | 25.0 ± 7.9 | 0.11 |
| **Lung function before exacerbation** |  |  |  |  |
| FEV<sub>1</sub> > 50% | 29 (25%) | 15 (21%) | 14 (33%) | 0.061 |
| FEV<sub>1</sub> 30 - 50% | 46 (40%) | 26 (37%) | 20 (47%) |  |
| FEV<sub>1</sub> < 30% | 39 (34%) | 30 (42%) | 9 (21%) |  |
| **Comorbidities** |  |  |  |  |
| Diabetes | 16 (13%) | 7 (9%) | 9 (19%) | 0.13 |
| Hypertension | 44 (36%) | 28 (37%) | 16 (33%) | 0.65 |
| Acute myocardial infarction | 33 (27%) | 17 (23%) | 16 (33%) | 0.19 |
| Atrial fibrillation | 20 (16%) | 6 (8%) | 14 (29%) | 0.002 |
| Emphysema | 40 (33%) | 27 (36%) | 13 (27%) | 0.30 |
| **Medications** |  |  |  |  |
| RAAS-inhibitors | 50 (41%) | 25 (33%) | 25 (52%) | 0.039 |
| Beta Blockers | 54 (44%) | 25 (33%) | 29 (60%) | 0.003 |
| Diuretics as regular medicine | 52 (42%) | 20 (27%) | 32 (67%) | <0.001 |
| Diuretics prior to LUS | 61 (50%) | 25 (33%) | 36 (75%) | <0.001 |
| **Baseline** |  |  |  |  |
| Respiration frequency (/min) | 27 ± 7 | 26 ± 6 | 28 ± 6 | 0.022 |
| Saturation (%) | 89 ± 7 | 89 ± 7 | 89 ± 6 | 0.89 |
| Systolic blood pressue (mmHg) | 139 ± 25 | 137 ± 23 | 141 ± 27 | 0.44 |
| Pulse (beats/minute) | 95 ± 18 | 97 ± 17 | 91 ± 20 | 0.067 |
| **Laboratory values** |  |  |  |  |
| CRP (ng/L) | 20 (7-60) | 21 (5-70) | 17 (11-47) | 0.99 |
| NT-proBNP (ng/L) | 455 (125-1247) | 125 (39-373) | 979 (490-2896) | <0.001 |
| High sensitive Troponine T (ng/L) | 30 (18-52) | 24 (14-41) | 45 (26-58) | <0.001 |
| Creatinine (umol/L) | 69 (58-92) | 67 (55-87) | 75 (59-114) | 0.030 |
| Hemoglobin (g/dL) | 13.6 ± 1.9 | 13.9 ± 1.7 | 13.1 ± 2.0 | 0.028 |
| Partial pressue oxygen (kPa) | 8.8 ± 3.4 | 9.2 ± 4.1 | 8.2 ± 1.7 | 0.098 |
| **Chest X-ray** |  |  |  |  |
| Congestion | 36 (30%) | 9 (13%) | 27 (56%) | <0.001 |
| Infiltrate | 29 (24%) | 14 (19%) | 15 (31%) | 0.14 |
| **Index hospitalization** |  |  |  |  |
| Length of stay (days) | 5 (3-7) | 4 (3-6) | 5 (3-10) | 0.15 |

CRP=C-reactive proteon, FEV<sub>1</sub>=forced expiratory volume in the 1<sup>st</sup> second, (n=114), HF=heart failure, hs-cTnT = high sensitive cardiac Troponine T (n=106), LUS=lung ultrasound, NT-proBNP (n=92), RAAS=renin-angiotensin system antagonist

**Table 2.** Events during 12 months follow up total, and by cut off for a positive LUS at ≥3 in ≥2 zones bilaterally

|  | **Negative**<br>**LUS** | **Positive**<br>**LUS** | **p-value** |
| --- | --- | --- | --- |
|  | ***N=107*** | ***N=16*** |  |
| Hospitalization (n, %) | 66 (62%) | 9 (56%) | - |
| Deaths | 15 (14%) | 2 (13%) | - |
| Composite | 81 (76%) | 11 (69%) | - |
| **Cox-regression** |  |  |  |
| Unadjusted | Ref | HR 0.98<br>(95% CI 0.52-1.84) | 0.94 |
| Adjusted for age and sex | Ref | HR 0.94<br>(95% CI 0.50-1.78) | 0.86 |
| Adjusted for age, sex and FEV<sub>1</sub> | Ref | HR 0.93<br>(95% CI 0.49-1.75) | 0.81 |

CI=confidence interval, HR=hazard ratio, FEV<sub>1</sub>=forced expiratory volume in the 1<sup>st</sup> second, LUS=lung ultrasound

Concurrent HF during the admission for AECOPD was present in 48 (39%) patients and 16 (13%) had coexisting HF but not considered clinically relevant for the current admission. A total of 47 (45%) of the patients adjudicated to have concurrent or co-existing HF did not have prior diagnosis of HF. Patients with concurrent HF were older, with less severe COPD, more prevalent AF, used more frequently RAAS-inhibitors, beta blockers and diuretics, had more frequently congestion on CXR and higher levels of cTnT, creatinine and NT-proBNP (median [IQR] 979 [490-2896 vs 125 [39-373 ng/L) (**Table 1**)

### Predictors for B-lines on lung ultrasound

Patients with positive LUS had comparable age, sex distribution, BMI, FEV_1_, comorbidity burden, and vital parameters to those with a negative LUS. (**Suppl. Table 1**). Patients with a positive vs negative LUS had more often presence of an infiltrate on CXR: 21 (20%) vs 8 (53%), p=0.005. The median level of NT-proBNP was 855 (130-2949) and 444 (120-1029) ng/L in patients with and without positive LUS, respectively (p=0.29).

The total number of B-lines across 8 zones was associated with older age, lower BMI, lower arterial partial pressure, diuretics use, higher CRP and the presence of infiltrates on CXR (**Suppl. Table 2**: **Figure 2**).

**Figure 2.**
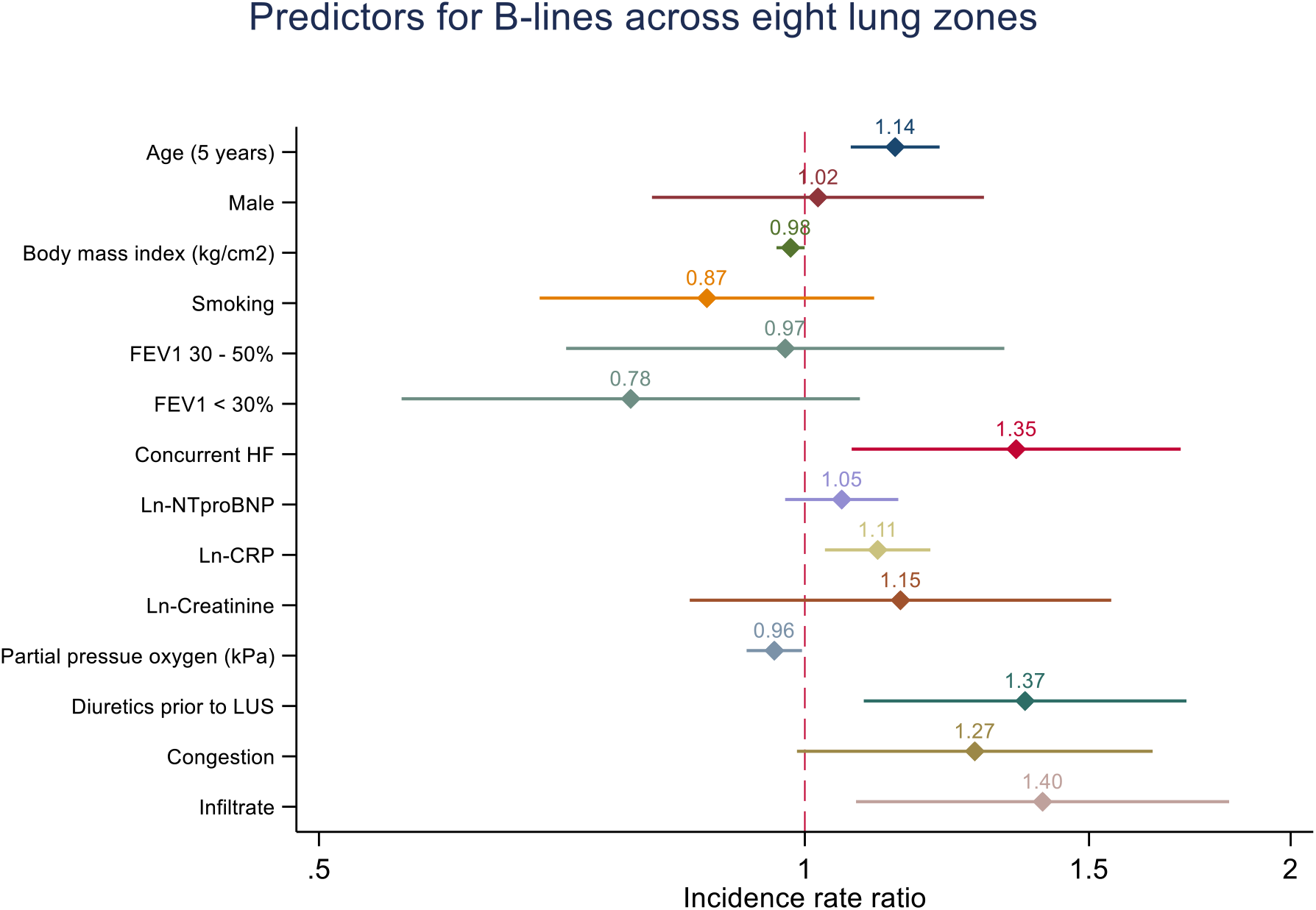
Clinical characteristics and findings in association with the total number of B-lines on LUS. Presented as a coefficient plot with incidence rate ratio and 95% confidence intervals. CRP=C-reative protein, FEV_1_=Forced expiratory volume during 1^st^ second, HF=heart failure, LUS=lung ultrasound, NTproBNP=N-terminal pro natriuretic peptide

**Figure 3.**
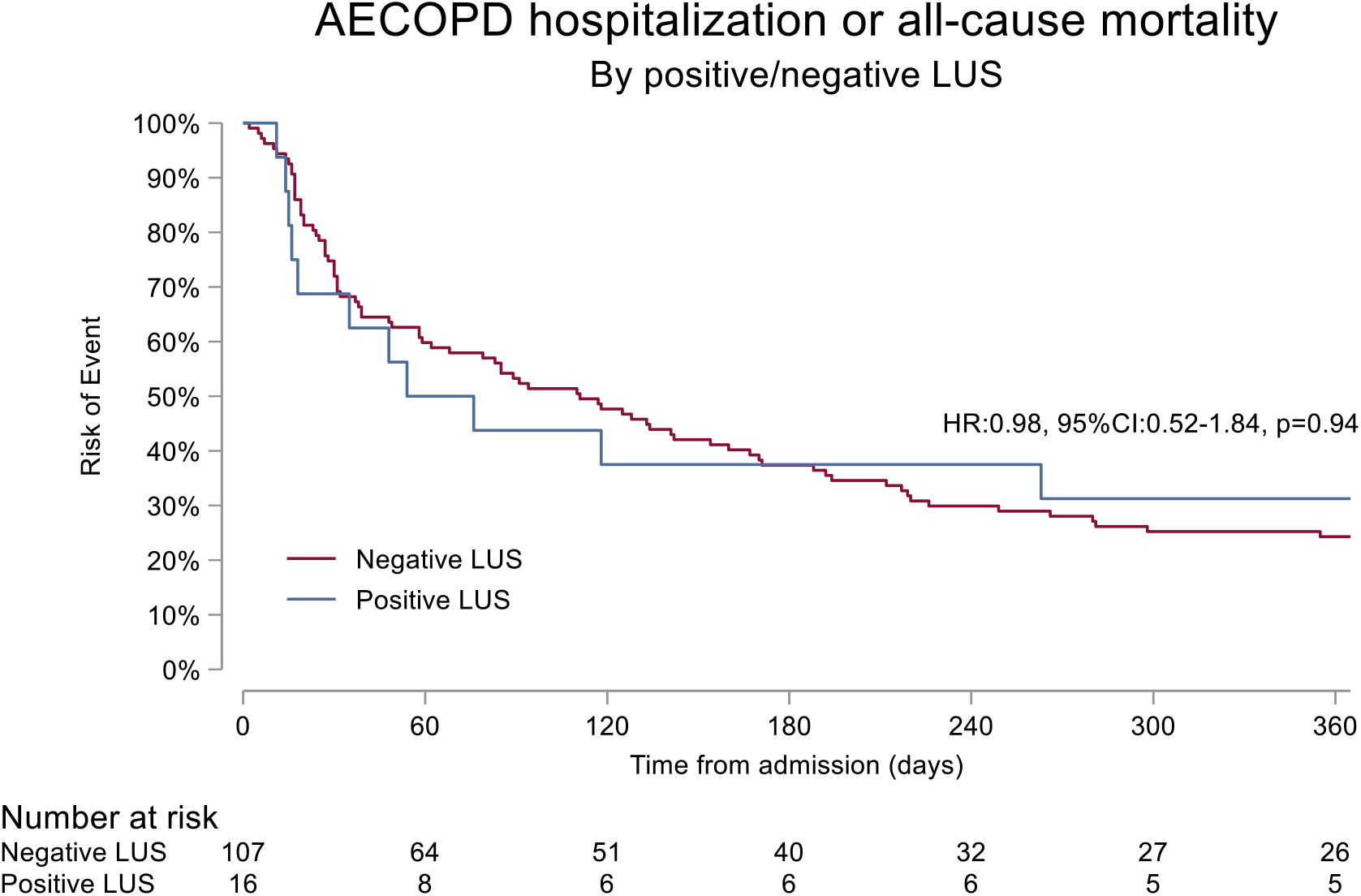
Survival plots for rehospitalization and all-cause mortality in patients with and without a positive LUS during the index admission, AECOPD=acute exacerbation of chronic obstructive pulmonary disorder, CI=confidence intervals, HR = hazard ratio, LUS=lung ultrasound

### Performance of LUS for detecting concurrent heart failure

The median number of B-lines was 10 (IQR 6–16) for patients with concurrent HF and 7 (IQR 5-12) for patients without concurrent HF (p=0.03). Concurrent acute HF was positively associated with the number of B-lines across 8 zones (ratio 1.35 [95% CI 1.07-1.71], p=0.012), but not after adjusting for age, BMI, CRP, diuretics use and infiltrate on CXR (p=0.18).

Sixteen patients (13%) had a positive LUS, and there was no difference between patients with and without concurrent HF: 8 (17%) vs 8 (11%), p=0.34. The ROC AUC, sensitivity and specificity for a positive LUS for correctly classifying concurrent HF was 0.53 (95% CI 0.47-0.59), 17% (95% CI 7-30%) and 89% (95% CI 80-95%), respectively. These measures were consistent in a sensitivity analysis excluding 16 patients with HF but not concurrent with AECOPD. We also obtained similar results in sensitivity analyses excluding patients with missing lung zones. Exploring ≥3 B-lines in one positive zone bilaterally as the definition of a positive LUS, the prevalence was 23 of 48 for patients with concurrent HF, and 20 of 75 for patients without concurrent HF (AUC=0.61).

### Association between findings from LUS and clinical outcomes

During the 12 months follow-up period after enrollment, there were in total 92 events (75 rehospitalizations for AECOPD and 17 deaths, **Table 3 and Figure 4**). Two (13%) patients in the positive LUS group and 15 (21%) in the negative LUS group died (p=0.87). Nine (56%) patients with positive LUS versus 66 (62%) with negative LUS were hospitalized for AECOPD (p=0.68). There were no significant association between positive LUS and time-to-event in the Cox proportional hazard regression analysis (HR 0.98 [95% CI 0.52-1.84], p=0.94), with consistent results after adjustments.

## Discussion

The main finding of our study is that bedside LUS is easily applicable in patients hospitalized with AECOPD but perform weakly as a diagnostic tool for the detection of concurrent HF. Furthermore, B-lines on LUS during hospitalization had no prognostic value for re-hospitalizations and all-cause mortality.

Our findings from patients with AECOPD contrast the promising evidence for LUS in managing patients with suspected acute HF and acute dyspnea ^9^. To our knowledge, there are few comparable studies with lung ultrasound in an AECOPD population. Based on prior studies and pathophysiological considerations in COPD, we hypothesized that LUS would detect concurrent HF in patients with AECOPD. There are some possible explanations for the contrasting results. First, the most common etiology for AECOPD is lower airway infection. We found consolidations on CXR to be the most robust clinical variable associated with B-lines. Thus, in AECOPD, lung parenchyma inflammation might disturb the expected association between pulmonary congestion and B-lines and decrease the diagnostic value of LUS in this population. In the subset of patients with follow-up LUS, we report that the number of B-lines does not diminish to day 2, which may support the latter explanation. A recent trial from intensive care patients reported poor ability of LUS to discriminate between pulmonary congestion and pneumonia.^22^ Second, AECOPD was the tentative diagnosis at admission in all patients. The HF severity might have been lower than in patients admitted to the emergency department with acute HF as the primary diagnosis. Indeed, the median number of B-lines was lower than comparable studies with acute HF patients.^23^ Third, we performed LUS on the first day in the medical ward. Many patients had already received diuretics during the first 24h, which may have decreased the pulmonary congestion and B-lines.^19^

The optimal approach to LUS imaging and B-line quantification is debated and our threshold for a positive LUS may be too strict.^11,24^ A recent methodological study with patients in the ED found that a 6-zone and 8-zone method, using ≥3 B-lines in one zone bilaterally as a cut-off, improved the diagnostic accuracy in patients with an unclear diagnosis of AHF, compared to several other thresholds (two positive zones bilateral threshold, ≥15 B-lines and ≥30 B-lines).^24^ A correlation between the bilateral presence of ≥3 B-lines in ≥1 zones and higher natriuretic peptide levels have been reported in an AECOPD-population.^25^ We found a similar trend in our study using ≥1 zones, but this was not statistically significant. Patients with concurrent HF had 35% more B-lines than patients with either non-concurrent HF or no HF.Prior studies suggest that using B-lines as a continuous parameter performs better with respect to diagnosis and prognosis compared to using dichotomized cut-offs.^7^ For example, a recent randomized trial in acute HF comparing LUS-guided therapy to standard care in acute HF failed to reach a pre-defined B-line threshold, despite a significant overall reduction in B-lines.^26^ Other studies in acute HF have found worse short-and long-term outcomes associated with higher B-lines.^13^ These results and the current study suggest that pulmonary congestion may be better depicted as a spectrum of B-lines rather than thresholds in patients with established COPD.

We and others have previously shown that cardiac comorbidity is associated with a poor outcome in patients with moderate or severe COPD.^27^ Subsequently, cardiac biomarkers such as natriuretic peptides and troponin seem to have independent prognostic value beyond COPD-related variables.^28,29^ B-lines on LUS have been associated with worse outcome in patients with AHF. However, we did not find any association between the presence of B-lines and the risk of readmission or death in patients with AECOPD. Considering that LUS associated more strongly with pulmonary infiltrates than concurrent HF in our study, this may not be surprising.

## Strengths and limitations

The strength of the study was that all patients underwent the same 8 zone protocol in semirecumbent positions, and the B-line readers were blinded clinically and temporally. The study also addresses the paucity of studies with lung ultrasound in the AECOPD population.

There are several significant limitations. First, our study lacks study-specific echocardiographic imaging variables and clinical classifications associated with increased cardiac filling pressures, recommended in the work-up of acute HF. We tried to overcome this limitation with HF adjudication by a CEC who had access to routine echocardiographic examinations, clinical examination reports etc. from the medical records. We used two different probes in our study, which may introduce bias. In the first period we used a linear probe with a limited depth (max 5.2 cm). The insufficient depth may, in theory, lead to the misinterpretation of pleural artifacts without clinical significance (Z-lines) as B-lines.

However, this limitation is more relevant in obese patients, and our patients were normal- or underweight. Although we did not measure skin-to-probe distance, we found only weak correlations between BMI and B-lines. No significant differences in the B-line count and relation to the outcome were detected comparing the two study periods. It is unclear whether the COVID-19 pandemic introduced bias, other than excluding a larger-than-usual portion of the COPD population. Of note, patients with COVID-19 were excluded per protocol. Lastly, information lost in a time-to-first as compared to recurrent event analysis may obscure the actual severity of B-line presence in AECOPD.

## Conclusion

Amongst patients hospitalized for AECOPD, B-lines are frequently observed on LUS. Concurrent acute HF was present in about one-third of patients and these patients had a higher number of B-lines. However, LUS with a threshold of a bilateral finding of ≥3 B-lines in two zones could not reliably detect concurrent HF. Infiltrates on CXR was the most important predictor of B-lines. Although assessment of B-lines by LUS is recommended by HF guidelines, our results suggest that it may be less useful for detecting HF in patients with AECOPD.

## Supporting information

Disclosures

## Data Availability

All data produced in the present study are available upon reasonable request to the authors

## References

1. Soriano JB, Abajobir AA, Abate KH, et al. Global, regional, and national deaths, prevalence, disability-adjusted life years, and years lived with disability for chronic obstructive pulmonary disease and asthma, 1990–2015: a systematic analysis for the Global Burden of Disease Study 2015. The Lancet Respiratory Medicine. 2017;5(9):691–706.

2. McDonagh TA, Metra M, Adamo M, et al. 2021 ESC Guidelines for the diagnosis and treatment of acute and chronic heart failure. Eur Heart J. 2021;42(36):3599–3726.

3. Maclay JD, MacNee W. Cardiovascular disease in COPD: mechanisms. Chest. 2013;143(3):798–807.

4. Cuthbert JJ, Kearsley JW, Kazmi S, et al. The impact of heart failure and chronic obstructive pulmonary disease on mortality in patients presenting with breathlessness. Clin Res Cardiol. 2019;108(2):185–193.

5. Volpicelli G, Cardinale L, Garofalo G, Veltri A. Usefulness of lung ultrasound in the bedside distinction between pulmonary edema and exacerbation of COPD. Emerg Radiol. 2008;15(3):145–151.

6. Prosen G, Klemen P, Strnad M, Grmec S. Combination of lung ultrasound (a comet-tail sign) and N-terminal pro-brain natriuretic peptide in differentiating acute heart failure from chronic obstructive pulmonary disease and asthma as cause of acute dyspnea in prehospital emergency setting. Crit Care. 2011;15(2):R114.

7. Platz E, Campbell RT, Claggett B, et al. Lung Ultrasound in Acute Heart Failure. JACC: Heart Failure. 2019;7(10):849–858.

8. Volpicelli G, Elbarbary M, Blaivas M, et al. International evidence-based recommendations for point-of-care lung ultrasound. Intensive Care Med. 2012;38(4):577–591.

9. Pivetta E, Goffi A, Nazerian P, et al. Lung ultrasound integrated with clinical assessment for the diagnosis of acute decompensated heart failure in the emergency department: a randomized controlled trial. Eur J Heart Fail. 2019;21(6):754–766.

10. Pivetta E, Goffi A, Lupia E, et al. Lung Ultrasound-Implemented Diagnosis of Acute Decompensated Heart Failure in the ED: A SIMEU Multicenter Study. Chest. 2015;148(1):202–210.

11. Laursen CB, Clive A, Hallifax R, et al. European Respiratory Society statement on thoracic ultrasound. Eur Respir J. 2021;57(3).

12. Platz E, Lewis EF, Uno H, et al. Detection and prognostic value of pulmonary congestion by lung ultrasound in ambulatory heart failure patients. Eur Heart J. 2016;37(15):1244–1251.

13. Platz E, Campbell RT, Claggett B, et al. Lung Ultrasound in Acute Heart Failure: Prevalence of Pulmonary Congestion and Short- and Long-Term Outcomes. JACC Heart Fail. 2019;7(10):849–858.

14. Cogliati C, Casazza G, Ceriani E, et al. Lung ultrasound and short-term prognosis in heart failure patients. Int J Cardiol. 2016;218:104–108.

15. Palazzuoli A, Ruocco G, Beltrami M, Nuti R, Cleland JG. Combined use of lung ultrasound, B-type natriuretic peptide, and echocardiography for outcome prediction in patients with acute HFrEF and HFpEF. Clin Res Cardiol. 2018;107(7):586–596.

16. McMurray JJ, Adamopoulos S, Anker SD, et al. ESC guidelines for the diagnosis and treatment of acute and chronic heart failure 2012: The Task Force for the Diagnosis and Treatment of Acute and Chronic Heart Failure 2012 of the European Society of Cardiology. Developed in collaboration with the Heart Failure Association (HFA) of the ESC. Eur J Heart Fail. 2012;14(8):803–869.

17. Zoneff ER, Baker K, Sweeny A, Keijzers G, Sanderson J, Watkins S. The prevalence of lung surface abnormalities in a healthy population as detected by a screening lung ultrasound protocol: Comparison between young and older volunteers. Australasian Journal of Ultrasound in Medicine. 2019;22(2):129–137.

18. Chiesa AM, Ciccarese F, Gardelli G, et al. Sonography of the normal lung: Comparison between young and elderly subjects. J Clin Ultrasound. 2015;43(4):230–234.

19. Martindale JL. Resolution of sonographic B-lines as a measure of pulmonary decongestion in acute heart failure. Am J Emerg Med. 2016;34(6):1129–1132.

20. Martindale JL, Noble VE, Liteplo A. Diagnosing pulmonary edema: lung ultrasound versus chest radiography. Eur J Emerg Med. 2013;20(5):356–360.

21. Brainin P, Claggett B, Lewis EF, et al. Body mass index and B-lines on lung ultrasonography in chronic and acute heart failure. ESC Heart Fail. 2020;7(3):1201–1209.

22. Bataille B, Riu B, Ferre F, et al. Integrated use of bedside lung ultrasound and echocardiography in acute respiratory failure: a prospective observational study in ICU. Chest. 2014;146(6):1586–1593.

23. Palazzuoli A, Evangelista I, Beltrami M, et al. Clinical, Laboratory and Lung Ultrasound Assessment of Congestion in Patients with Acute Heart Failure. J Clin Med. 2022;11(6).

24. Buessler A, Chouihed T, Duarte K, et al. Accuracy of Several Lung Ultrasound Methods for the Diagnosis of Acute Heart Failure in the ED: A Multicenter Prospective Study. Chest. 2020;157(1):99–110.

25. Sriram KB, Singh M. Lung ultrasound B-lines in exacerbations of chronic obstructive pulmonary disease. Intern Med J. 2017;47(3):324–327.

26. Pang PS, Russell FM, Ehrman R, et al. Lung Ultrasound-Guided Emergency Department Management of Acute Heart Failure (BLUSHED-AHF): A Randomized Controlled Pilot Trial. JACC Heart Fail. 2021;9(9):638–648.

27. Sin DD, Anthonisen NR, Soriano JB, Agusti AG. Mortality in COPD: Role of comorbidities. Eur Respir J. 2006;28(6):1245–1257.

28. Hoiseth AD, Brynildsen J, Hagve TA, et al. The influence of heart failure co-morbidity on high-sensitivity troponin T levels in COPD exacerbation in a prospective cohort study: data from the Akershus cardiac examination (ACE) 2 study. Biomarkers. 2016;21(2):173–179.

29. Hoiseth AD, Omland T, Hagve TA, Brekke PH, Soyseth V. NT-proBNP independently predicts long term mortality after acute exacerbation of COPD - a prospective cohort study. Respir Res. 2012;13:97.

