## Supplementary Material for "Lung ultrasound to assess pulmonary congestion in patients with acute exacerbation of COPD - a feasibility study"

### Supplemental Material

### Imputation of B-lines

In patients with missing B-line data in ≤2 out of 4 zones (n=7), we imputed B-line data from anatomically adjacent zones: Zones 1 and 2, zones 3 and 4, zones 5 and 6, zones 7 and 8.

| | **Zones 1 & 2** | **Zones 3 & 4** | **Zones 5 & 6** | **Zones 7 & 8** |
|---|---|---|---|---|
| Correlation between adjacent zones (rho, P) | 0.32 (p<0.001) | 0.14 (p=0.13) | 0.28 (p=0.002) | 0.13 (p=0.154) |
| Imputed zones (n) | n=1 | n=1 | n=2 | n=3 |

**Supplementary Table S1. Baseline characteristics of total population and according to lung ultrasound**

| | Total | Negative LUS | Positive LUS | p-value |
|---|---|---|---|---|
| | ***N=123*** | ***N=107*** | ***N=16*** | |
| Age (years) | 75 ± 9 | 74 ± 9 | 75 ± 9 | 0.74 |
| Male | 57 (46%) | 50 (47%) | 7 (44%) | 0.82 |
| Smoking | 52 (42%) | 43 (40%) | 9 (56%) | 0.23 |
| Body mass index (kg/cm2) | 23.8 ± 6.4 | 23.8 ± 6.4 | 23.7 ± 6.3 | 0.95 |
| $FEV_1$ | | | | |
| $FEV_1$ > 50% | 29 (25%) | 24 (24%) | 5 (31%) | 0.85 |
| $FEV_1$ 30 - 50% | 46 (40%) | 40 (41%) | 6 (38%) | |
| $FEV_1$ < 30% | 39 (34%) | 34 (35%) | 5 (31%) | |
| Diabetes | 16 (13%) | 13 (12%) | 3 (19%) | 0.46 |
| Hypertension | 44 (36%) | 36 (34%) | 8 (50%) | 0.20 |
| Acute myocardial infarction | 33 (27%) | 29 (27%) | 4 (25%) | 0.86 |
| Atrial fibrillation | 20 (16%) | 18 (17%) | 2 (13%) | 0.66 |
| Diuretics pre-admission | 52 (42%) | 44 (41%) | 8 (50%) | 0.50 |
| Diuretics prior to LUS | 61 (50%) | 55 (51%) | 6 (38%) | 0.30 |
| Respiration frequency (breaths/minute) | 27 ± 7 | 27 ± 7 | 27 ± 7 | 0.96 |
| Peripheral $O_2$ saturation (%) | 89 ± 7 | 89 ± 7 | 87 ± 7 | 0.21 |
| Systolic blood pressue (mmHg) | 139 ± 25 | 138 ± 24 | 144 ± 28 | 0.33 |
| Pulse (beats/minute) | 95 ± 18 | 95 ± 19 | 93 ± 13 | 0.67 |
| C-reactive protein (ng/L) | 20 (7-60) | 21 (6-60) | 16 (10-71) | 0.84 |
| NT-proBNP (ng/L) | 455 (125-1247) | 444 (120-1029) | 855 (130-2949) | 0.29 |
| High sensitive Troponin T (ng/L) | 30 (18-52) | 31 (17-52) | 28 (20-45) | 0.66 |
| Creatinine (umol/L) | 69 (58-92) | 69 (57-98) | 71 (59-89) | 0.72 |
| Hemoglobin (g/dL) | 13.6 ± 1.9 | 13.6 ± 1.9 | 13.7 ± 1.5 | 0.74 |
| Arterial $O_2$ (kPa) | 8.8 ± 3.4 | 8.9 ± 3.6 | 8.3 ± 1.5 | 0.54 |
| Chest X-ray - congestion | 36 (30%) | 32 (30%) | 4 (27%) | 0.76 |
| Chest X-ray - infiltrate | 29 (24%) | 21 (20%) | 8 (53%) | 0.005 |
| Length of stay (days) | 5 (3-7) | 5 (3-8) | 4 (3-6) | 0.41 |

$FEV_1$=forced expiratory volume in the 1$^{st}$ second, (n=114), LUS=lung ultrasound, NTproBNP=N-terminal pro-brain natriuretic peptide

**Supplementary Table 2. Predictors of the total number of B-lines on LUS. Analyzed by negative binomial regression with total B-lines number as dependent variable[1]**

| **Independent variable** | **IRR (95% CI)** | **p-value** |
|---|---|---|
| Age (per 5 years) | 1.14 (1.07-1.21) | <0.001 |
| Male | 1.02 ( 0.80-1.29) | 0.88 |
| BMI (per unit) | 0.98 (0.69-1.10) | 0.047 |
| Smoking | 0.87 (-0.38- 0.10) | 0.25 |
| Emphysema | 1.12 (0.87-1.43) | 0.39 |
| $FEV_1$ 30-50% | 0.97 (0.71-1.33) | 0.86 |
| $FEV_1$ <30% | 0.78 (0.56-1.08) | 0.14 |
| Concurrent acute HF | 1.40 (1.11-1.76) | 0.004 |
| NTproBNP (per ln increase) | 1.05 (0.97-1.14) | 0.20 |
| CRP (per ln increase) | 1.11 (1.03-1.20) | 0.007 |
| Creatinine (per umol/L) | 1.15 (0.85-1.55) | 0.37 |
| Arterial pressure O2 (kPa) | 0.96 (0.93- 0.99) | 0.005 |
| Diuretics given prior to LUS | 1.37 (1.09-1.72) | 0.007 |
| Diuretics newly initiated | 1.44 (1.04-1.99) | 0.029 |
| Non-invasive ventilation | 0.88 (0.68-1.14) | 0.34 |
| Chest X-ray - congestion | 1.27 (0.99-1.64) | 0.061 |
| Chest X-ray - infiltrate | 1.40, (1.08-1.83) | 0.012 |

BMI=body mass index, CRP=C-reactive protein, $FEV_1$=forced expiratory volume in the 1st second, (n=114), HF=heart failure, IRR=incidence-rate ratios, LUS=lung ultrasound, NTproBNP=N-terminal pro-brain natriuretic peptide

[1]one unit increase in the independent variable leads to % change in sum of B-lines in 8 zones.
