## Supplementary material for "Lung ultrasound to assess pulmonary congestion in patients with acute exacerbation of COPD - a feasibility study": Disclosures

### ICMJE DISCLOSURE FORM

**Date:** 7/18/2022

**Your Name:** Fride Uthaug Reite

**Manuscript Number (if known):** Click or tap here to enter text.

In the interest of transparency, we ask you to disclose all relationships/activities/interests listed below that are related to the content of your manuscript. "Related" means any relation with for-profit or not-for-profit third parties whose interests may be affected by the content of the manuscript. Disclosure represents a commitment to transparency and does not necessarily indicate a bias. If you are in doubt about whether to list a relationship/activity/interest, it is preferable that you do so.

The author's relationships/activities/interests should be defined broadly. For example, if your manuscript pertains to the epidemiology of hypertension, you should declare all relationships with manufacturers of antihypertensive medication, even if that medication is not mentioned in the manuscript.

In item #1 below, report all support for the work reported in this manuscript without time limit. For all other items, the time frame for disclosure is the past 36 months.

| | | Name all entities with whom you have this relationship or indicate none (add rows as needed) | Specifications/Comments (e.g., if payments were made to you or to your institution) |
|---|---|---|---|
| | | **Time frame: Since the initial planning of the work** | |
| 1 | All support for the present manuscript (e.g., funding, provision of study materials, medical writing, article processing charges, etc.) **No time limit for this item.** | ☐ None | |
| | | Norwegian Society of Pulmonary Physicians | Grant sponsored by Aztra Zeneca, paid to the institution.<br>The sponsor had no input or contribution in the development of the research and manuscript the tab key to add additional rows |
| | | **Time frame: past 36 months** | |
| **2** | Grants or contracts from any entity (if not indicated in item #1 above). | ☒ **None** | |
| **3** | Royalties or licenses | ☒ **None** | |

| | | Name all entities with whom you have this relationship or indicate none (add rows as needed) | Specifications/Comments (e.g., if payments were made to you or to your institution) |
|---|---|---|---|
| **4** | Consulting fees | ☒ **None** | |
| **5** | Payment or honoraria for lectures, presentations, speakers bureaus, manuscript writing or educational events | ☒ **None** | |
| **6** | Payment for expert testimony | ☒ **None** | |
| **7** | Support for attending meetings and/or travel | ☒ **None** | |
| **8** | Patents planned, issued or pending | ☒ **None** | |
| **9** | Participation on a Data Safety Monitoring Board or Advisory Board | ☒ **None** | |
| **10** | Leadership or fiduciary role in other board, society, committee or advocacy group, paid or unpaid | ☒ **None** | |

| | | Name all entities with whom you have this relationship or indicate none (add rows as needed) | Specifications/Comments (e.g., if payments were made to you or to your institution) |
|---|---|---|---|
| **11** | Stock or stock options | ☒ **None** | |
| **12** | Receipt of equipment, materials, drugs, medical writing, gifts or other services | ☒ **None** | |
| **13** | Other financial or non-financial interests | ☒ **None** | |

**Please place an "X" next to the following statement to indicate your agreement:**

☒ I certify that I have answered every question and have not altered the wording of any of the questions on this form.

### ICMJE DISCLOSURE FORM

| | |
|---|---|
| **Date:** | 7/18/2022 |
| **Your Name:** | Rahul Bhatnagar |
| **Manuscript Title:** | Lung ultrasound to assess pulmonary congestion in patients with acute exacerbation of COPD – a feasibility study |
| **Manuscript Number (if known):** | Click or tap here to enter text. |

In the interest of transparency, we ask you to disclose all relationships/activities/interests listed below that are related to the content of your manuscript. "Related" means any relation with for-profit or not-for-profit third parties whose interests may be affected by the content of the manuscript. Disclosure represents a commitment to transparency and does not necessarily indicate a bias. If you are in doubt about whether to list a relationship/activity/interest, it is preferable that you do so.

The author's relationships/activities/interests should be defined broadly. For example, if your manuscript pertains to the epidemiology of hypertension, you should declare all relationships with manufacturers of antihypertensive medication, even if that medication is not mentioned in the manuscript.

In item #1 below, report all support for the work reported in this manuscript without time limit. For all other items, the time frame for disclosure is the past 36 months.

| | | **Name all entities with whom you have this relationship or indicate none (add rows as needed)** | **Specifications/Comments (e.g., if payments were made to you or to your institution)** |
|---|---|---|---|
| | | Time frame: Since the initial planning of the work | |
| 1 | All support for the present manuscript (e.g., funding, provision of study materials, medical writing, article processing charges, etc.) **No time limit for this item.** | ☒ None | |
| | | Time frame: past 36 months | |
| **2** | Grants or contracts from any entity (if not indicated in item #1 above). | ☒ **None** | |
| **3** | Royalties or licenses | ☒ **None** | |

| | | Name all entities with whom you have this relationship or indicate none (add rows as needed) | Specifications/Comments (e.g., if payments were made to you or to your institution) |
|---|---|---|---|
| **4** | Consulting fees | ☒ **None** | |
| **5** | Payment or honoraria for lectures, presentations, speakers bureaus, manuscript writing or educational events | ☒ **None** | |
| **6** | Payment for expert testimony | ☒ **None** | |
| **7** | Support for attending meetings and/or travel | ☒ **None** | |
| **8** | Patents planned, issued or pending | ☒ **None** | |
| **9** | Participation on a Data Safety Monitoring Board or Advisory Board | ☒ **None** | |
| **10** | Leadership or fiduciary role in other board, society, committee or advocacy group, paid or unpaid | ☒ **None** | |

| | | Name all entities with whom you have this relationship or indicate none (add rows as needed) | Specifications/Comments (e.g., if payments were made to you or to your institution) |
|---|---|---|---|
| **11** | Stock or stock options | ☒ **None** | |
| **12** | Receipt of equipment, materials, drugs, medical writing, gifts or other services | ☒ **None** | |
| **13** | Other financial or non-financial interests | ☒ **None** | |

**Please place an "X" next to the following statement to indicate your agreement:**

☒ I certify that I have answered every question and have not altered the wording of any of the questions on this form.

### ICMJE DISCLOSURE FORM

**Date:** 7/18/2022

**Your Name:** Peder Langeland Myhre

**Manuscript Title:** Lung ultrasound to assess pulmonary congestion in patients with acute exacerbation of COPD – a feasibility study

**Manuscript Number (if known):** Click or tap here to enter text.

In the interest of transparency, we ask you to disclose all relationships/activities/interests listed below that are related to the content of your manuscript. "Related" means any relation with for-profit or not-for-profit third parties whose interests may be affected by the content of the manuscript. Disclosure represents a commitment to transparency and does not necessarily indicate a bias. If you are in doubt about whether to list a relationship/activity/interest, it is preferable that you do so.

The author's relationships/activities/interests should be defined broadly. For example, if your manuscript pertains to the epidemiology of hypertension, you should declare all relationships with manufacturers of antihypertensive medication, even if that medication is not mentioned in the manuscript.

In item #1 below, report all support for the work reported in this manuscript without time limit. For all other items, the time frame for disclosure is the past 36 months.

| | | Name all entities with whom you have this relationship or indicate none (add rows as needed) | Specifications/Comments (e.g., if payments were made to you or to your institution) |
|---|---|---|---|
| Time frame: Since the initial planning of the work | | | |
| 1 | All support for the present manuscript (e.g., funding, provision of study materials, medical writing, article processing charges, etc.) **No time limit for this item.** | ☒ None | |
| Time frame: past 36 months | | | |
| 2 | Grants or contracts from any entity (if not indicated in item #1 above). | ☐ **None** | |
| | | AstraZeneca | To Akershus University Hospital |
| 3 | Royalties or licenses | ☒ **None** | |

| | | Name all entities with whom you have this relationship or indicate none (add rows as needed) | Specifications/Comments (e.g., if payments were made to you or to your institution) |
|---|---|---|---|
| **4** | Consulting fees | ☐ **None** | |
| | | Astra Zeneca | Consulting fees |
| | | Boehringer-Ingelheim | Consulting fees |
| | | Novartis | Consulting fees |
| | | Novo Nordisk | Consulting fees |
| **5** | Payment or honoraria for lectures, presentations, speakers bureaus, manuscript writing or educational events | ☐ **None** | |
| | | Amarin | Honoraria for lectures |
| | | AmGen | Honoraria for lectures |
| | | Astra Zeneca | Honoraria for lectures |
| | | Bayer | Honoraria for lectures |
| | | Boehringer-Ingelheim | Honoraria for lectures |
| | | Novartis | Honoraria for lectures |
| | | Novo Nordisk | Honoraria for lectures |
| **6** | Payment for expert testimony | ☒ **None** | |
| **7** | Support for attending meetings and/or travel | ☒ **None** | |
| **8** | Patents planned, issued or pending | ☒ **None** | |
| **9** | Participation on a Data Safety Monitoring Board or Advisory Board | ☐ **None** | |
| | | Amarin | Advisory Boards |
| | | Astra Zeneca | Advisory Boards |
| | | Boehringer-Ingelheim | Advisory Boards |
| | | Novartis | Advisory Boards |
| | | Novo Nordisk | Advisory Boards |
| **10** | Leadership or fiduciary role in other board, society, committee or advocacy group, paid or unpaid | ☒ **None** | |

| | | Name all entities with whom you have this relationship or indicate none (add rows as needed) | Specifications/Comments (e.g., if payments were made to you or to your institution) |
|---|---|---|---|
| **11** | Stock or stock options | ☒ **None** | |
| **12** | Receipt of equipment, materials, drugs, medical writing, gifts or other services | ☒ **None** | |
| **13** | Other financial or non-financial interests | ☒ **None** | |

**Please place an "X" next to the following statement to indicate your agreement:**

☒ I certify that I have answered every question and have not altered the wording of any of the questions on this form.

### ICMJE DISCLOSURE FORM

| | |
|---|---|
| **Date:** | 7/18/2022 |
| **Your Name:** | Tarjei Øvrebotten |
| **Manuscript Title:** | Lung ultrasound to assess pulmonary congestion in patients with acute exacerbation of COPD – a feasibility study |
| **Manuscript Number (if known):** | Click or tap here to enter text. |

In the interest of transparency, we ask you to disclose all relationships/activities/interests listed below that are related to the content of your manuscript. "Related" means any relation with for-profit or not-for-profit third parties whose interests may be affected by the content of the manuscript. Disclosure represents a commitment to transparency and does not necessarily indicate a bias. If you are in doubt about whether to list a relationship/activity/interest, it is preferable that you do so.

The author's relationships/activities/interests should be defined broadly. For example, if your manuscript pertains to the epidemiology of hypertension, you should declare all relationships with manufacturers of antihypertensive medication, even if that medication is not mentioned in the manuscript.

In item #1 below, report all support for the work reported in this manuscript without time limit. For all other items, the time frame for disclosure is the past 36 months.

| | | **Name all entities with whom you have this relationship or indicate none (add rows as needed)** | **Specifications/Comments (e.g., if payments were made to you or to your institution)** |
|---|---|---|---|
| **Time frame: Since the initial planning of the work** | | | |
| 1 | All support for the present manuscript (e.g., funding, provision of study materials, medical writing, article processing charges, etc.)<br>No time limit for this item. | ☒ None | |
| **Time frame: past 36 months** | | | |
| **2** | Grants or contracts from any entity (if not indicated in item #1 above). | ☒ **None** | |
| **3** | Royalties or licenses | ☒ **None** | |

| | | Name all entities with whom you have this relationship or indicate none (add rows as needed) | Specifications/Comments (e.g., if payments were made to you or to your institution) |
|---|---|---|---|
| **4** | Consulting fees | ☒ **None** | |
| **5** | Payment or honoraria for lectures, presentations, speakers bureaus, manuscript writing or educational events | ☒ **None** | |
| **6** | Payment for expert testimony | ☒ **None** | |
| **7** | Support for attending meetings and/or travel | ☒ **None** | |
| **8** | Patents planned, issued or pending | ☒ **None** | |
| **9** | Participation on a Data Safety Monitoring Board or Advisory Board | ☒ **None** | |
| **10** | Leadership or fiduciary role in other board, society, committee or advocacy group, paid or unpaid | ☒ **None** | |

| | | Name all entities with whom you have this relationship or indicate none (add rows as needed) | Specifications/Comments (e.g., if payments were made to you or to your institution) |
|---|---|---|---|
| **11** | Stock or stock options | ☒ **None** | |
| **12** | Receipt of equipment, materials, drugs, medical writing, gifts or other services | ☒ **None** | |
| **13** | Other financial or non-financial interests | ☒ **None** | |

**Please place an "X" next to the following statement to indicate your agreement:**

☒ I certify that I have answered every question and have not altered the wording of any of the questions on this form.

### ICMJE DISCLOSURE FORM

**Date:** 7/18/2022

**Your Name:** Øyvind Johannessen

**Manuscript Title:** Lung ultrasound to assess pulmonary congestion in patients with acute exacerbation of COPD – a feasibility study

**Manuscript Number (if known):** Click or tap here to enter text.

In the interest of transparency, we ask you to disclose all relationships/activities/interests listed below that are related to the content of your manuscript. “Related” means any relation with for-profit or not-for-profit third parties whose interests may be affected by the content of the manuscript. Disclosure represents a commitment to transparency and does not necessarily indicate a bias. If you are in doubt about whether to list a relationship/activity/interest, it is preferable that you do so.

The author’s relationships/activities/interests should be defined broadly. For example, if your manuscript pertains to the epidemiology of hypertension, you should declare all relationships with manufacturers of antihypertensive medication, even if that medication is not mentioned in the manuscript.

In item #1 below, report all support for the work reported in this manuscript without time limit. For all other items, the time frame for disclosure is the past 36 months.

| | | **Name all entities with whom you have this relationship or indicate none (add rows as needed)** | **Specifications/Comments (e.g., if payments were made to you or to your institution)** |
|---|---|---|---|
| | | **Time frame: Since the initial planning of the work** | |
| 1 | All support for the present manuscript (e.g., funding, provision of study materials, medical writing, article processing charges, etc.) **No time limit for this item.** | ☐ None | |
| | | Norwegian Society of Pulmonary Physicians | Grant paid to the author<br>The sponsor had no input or contribution in the development of the research and manuscript. |
| | | The Norwegian society for Ultrasound in general practice | Grant paid to the author<br>The sponsor had no input or contribution in the development of the research and manuscript. |
| | | Norwegian Society of Pulmonary Physicians | Grant sponsored by Aztra Zenaca, paid to the institution.<br>The sponsor had no input or contribution in the development of the research and manuscript.C the tab key to add additional rows |
| | | **Time frame: past 36 months** | |
| 2 | Grants or contracts from any entity (if not indicated in item #1 above). | ☒ **None** | |

| | | Name all entities with whom you have this relationship or indicate none (add rows as needed) | Specifications/Comments (e.g., if payments were made to you or to your institution) |
|---|---|---|---|
| **3** | Royalties or licenses | ☒ **None** | |
| **4** | Consulting fees | ☒ **None** | |
| **5** | Payment or honoraria for lectures, presentations, speakers bureaus, manuscript writing or educational events | ☒ **None** | |
| **6** | Payment for expert testimony | ☒ **None** | |
| **7** | Support for attending meetings and/or travel | ☒ **None** | |
| **8** | Patents planned, issued or pending | ☒ **None** | |
| **9** | Participation on a Data Safety Monitoring Board or Advisory Board | ☒ **None** | |
| **10** | Leadership or fiduciary role in other board, | ☒ **None** | |

| | | Name all entities with whom you have this relationship or indicate none (add rows as needed) | Specifications/Comments (e.g., if payments were made to you or to your institution) |
|---|---|---|---|
| | society, committee or advocacy group, paid or unpaid | | |
| **11** | Stock or stock options | ☒ **None** | |
| **12** | Receipt of equipment, materials, drugs, medical writing, gifts or other services | ☒ **None** | |
| **13** | Other financial or non-financial interests | ☒ **None** | |

**Please place an "X" next to the following statement to indicate your agreement:**

☒ I certify that I have answered every question and have not altered the wording of any of the questions on this form.

### ICMJE DISCLOSURE FORM

**Date:** 7/18/2022

**Your Name:** Gunnar Einvik

**Manuscript Title:** Lung ultrasound to assess pulmonary congestion in patients with acute exacerbation of COPD – a feasibility study

**Manuscript Number (if known):** Click or tap here to enter text.

In the interest of transparency, we ask you to disclose all relationships/activities/interests listed below that are related to the content of your manuscript. "Related" means any relation with for-profit or not-for-profit third parties whose interests may be affected by the content of the manuscript. Disclosure represents a commitment to transparency and does not necessarily indicate a bias. If you are in doubt about whether to list a relationship/activity/interest, it is preferable that you do so.

The author's relationships/activities/interests should be defined broadly. For example, if your manuscript pertains to the epidemiology of hypertension, you should declare all relationships with manufacturers of antihypertensive medication, even if that medication is not mentioned in the manuscript.

In item #1 below, report all support for the work reported in this manuscript without time limit. For all other items, the time frame for disclosure is the past 36 months.

| | | Name all entities with whom you have this relationship or indicate none (add rows as needed) | Specifications/Comments (e.g., if payments were made to you or to your institution) |
|---|---|---|---|
| Time frame: Since the initial planning of the work | | | |
| 1 | All support for the present manuscript (e.g., funding, provision of study materials, medical writing, article processing charges, etc.) **No time limit for this item.** | ☐ None | |
| | | Norwegian Society of Pulmonary Physicians | Grant sponsored by Aztra Zenaca, paid to the institution.<br>The sponsor had no input or contribution in the development of the research and manuscript |
| Time frame: past 36 months | | | |
| **2** | Grants or contracts from any entity (if not indicated in item #1 above). | ☒ **None** | |
| **3** | Royalties or licenses | ☒ **None** | |

| | | Name all entities with whom you have this relationship or indicate none (add rows as needed) | Specifications/Comments (e.g., if payments were made to you or to your institution) |
|---|---|---|---|
| **4** | Consulting fees | ☒ **None** | |
| | | Astra Zeneca | Advisory board |
| **5** | Payment or honoraria for lectures, presentations, speakers bureaus, manuscript writing or educational events | ☒ **None** | |
| **6** | Payment for expert testimony | ☒ **None** | |
| **7** | Support for attending meetings and/or travel | ☒ **None** | |
| **8** | Patents planned, issued or pending | ☒ **None** | |
| **9** | Participation on a Data Safety Monitoring Board or Advisory Board | ☒ **None** | |
| **10** | Leadership or fiduciary role in other board, society, committee or advocacy group, paid or unpaid | ☒ **None** | |

| | | Name all entities with whom you have this relationship or indicate none (add rows as needed) | Specifications/Comments (e.g., if payments were made to you or to your institution) |
|---|---|---|---|
| **11** | Stock or stock options | ☒ **None** | |
| **12** | Receipt of equipment, materials, drugs, medical writing, gifts or other services | ☒ **None** | |
| **13** | Other financial or non-financial interests | ☒ **None** | |

**Please place an "X" next to the following statement to indicate your agreement:**

☒ I certify that I have answered every question and have not altered the wording of any of the questions on this form.
